# Detection of human brain cancers using genomic and immune cell characterization of cerebrospinal fluid through CSF-BAM

**DOI:** 10.1101/2024.12.02.24318303

**Authors:** Alexander H. Pearlman, Yuxuan Wang, Anita Kalluri, Megan Parker, Joshua D Cohen, Jonathan Dudley, Jordina Rincon-Torroella, Yuanxuan Xia, Ryan Gensler, Melanie Alfonzo Horwitz, John Theodore, Lisa Dobbyn, Maria Popoli, Janine Ptak, Natalie Silliman, Kathy Judge, Mari Groves, Christopher M. Jackson, Eric M. Jackson, George I. Jallo, Michael Lim, Mark Luciano, Debraj Mukherjee, Jarushka Naidoo, Sima Rozati, Cole H. Sterling, Jon Weingart, Carl Koschmann, Alireza Mansouri, Michael Glantz, David Kamson, Karisa C. Schreck, Carlos A. Pardo, Matthias Holdhoff, Suman Paul, Kenneth W. Kinzler, Nickolas Papadopoulos, Bert Vogelstein, Christopher Douville, Chetan Bettegowda

## Abstract

Patients who have radiographically detectable lesions in their brain or other symptoms compatible with brain tumors pose challenges for diagnosis. The only definitive way to diagnose such patients is through brain biopsy, an obviously invasive and dangerous procedure. Here we present a new workflow termed “CSF-BAM” that simultaneously identifies ***<u>B</u>*** cell or T cell receptor rearrangements, ***<u>A</u>***neuploidy, and ***<u>M</u>****utations* using PCR-mediated amplification of both strands of the DNA from CSF samples. We first describe the details of the molecular genetic assessments and then establish thresholds for positivity using training sets of libraries from patients with or without cancer. We then applied CSF-BAM to an independent set of 206 DNA samples from patients with common, aggressive cancer types as well as other forms of brain cancers. Among the 126 samples from patients with the most common aggressive cancer types (high grade gliomas, medulloblastomas, or metastatic cancers to the brain), the sensitivity of detection was >81%. None of 33 CSF-BAM assays (100% specificity, 90% to 100% credible interval) were positive in CSF samples from patients without brain cancers. The sensitivity of CSF-BAM was considerably higher than that achieved with cytology. CSF-BAM provides an integrated multi-analyte approach to identify neoplasia in the central nervous system, provides information about the immune environment in patients with or without cancer, and has the potential to inform the subsequent management of such patients.

**Statement of significance:** There is a paucity of technologies beyond surgical biopsy that can accurately diagnose central nervous system neoplasms. We developed a novel, sensitive and highly specific assay that can detect brain cancers by comprehensively identifying somatic mutations, chromosomal copy number changes, and adaptive immunoreceptor repertoires from samples of cerebrospinal fluid.

## Introduction

Brain cancers represent a heterogenous but highly aggressive class of neoplasms. They can be broadly categorized as primary or metastatic. Glioblastoma and medulloblastoma represent the most common types of primary brain cancers in adults and children respectively^1–3^. Brain metastases occur in over 200,000 individuals in the United States every year and can be classified as parenchymal, which occur in the substance of the brain and are more common, or leptomeningeal, which occur in the lining of the brain in up to 10% of all cancer patients^4,5^. Lung, breast, colon, melanoma, and renal cancers represent the most common cancer types to metastasize to the brain. Despite aggressive multi-modality treatments, both primary and metastatic brain cancers are associated with abysmal long-term survival, with most forms of brain cancer being incurable^6–9^.

The primary and almost exclusive methodology to diagnose brain cancers remains neurosurgical biopsy, which has significant inherent risks and expense. A brain biopsy is unlike tissue sampling in any other organ. It typically requires general anesthesia with inpatient hospitalization, and it carries a 5-10% risk for neurological decline, a 1% risk for catastrophic hemorrhage, a 5-10% risk for a non-diagnostic result and is susceptible to sampling bias as only a very small fragment of the tumor is extracted^10–16^. In select cases, such as CNS lymphoma or leptomeningeal disease (LMD), CSF cytology can aid in diagnosis but the sensitivity is relatively low^17–21^. To date, about 1 in 4 patients with LMD are diagnosed based on imaging or clinical findings while having negative cytology^22^. In addition, outside of tissue sampling, there is no methodology to understand the molecular and cellular composition of CNS cancers. However, it is becoming increasingly evident that understanding the genetic and epigenetic drivers of CNS neoplasms is essential for establishing a diagnosis for both primary and metastatic brain cancers, enabling prognostication and for informing therapeutic decision making^2,6,7,23–25^. The primary diagnostic criteria based on the WHO 2021 guidelines incorporate chromosomal copy number alterations and disease defining somatic mutations^26^. Interrogating CSF through liquid biopsies, which has demonstrated superior performance compared to evaluating peripheral blood, represents a promising means of accessing such genetic and epigenetic information for CNS neoplasms without the need for potentially morbid tissue biopsy^27–36^. Additionally, improved CSF liquid biopsy will provide essential information about response and resistance to investigational agents in clinical trials, particularly since repeat issue sampling is not commonly performed^37^.

In recent years, there has also been a burgeoning understanding of the central relationship between the immune microenvironment and susceptibility to immune modulating anti-cancer therapies^38–41^. For example, the advent of immune therapies such as checkpoint inhibitors and CAR T cell approaches has increased the need for understanding the relationships within immune cell populations associated with CNS cancers^42–45^. In addition, some of the rare but challenging primary brain cancers are primary central nervous system lymphomas (PCNSL), which are B cell lymphomas thought to originate in the brain^46,47^. Understanding the cancerous B cell clones and their molecular phenotype has important biological and therapeutic ramifications^48–52^. Similarly, CSF is often obtained in patients with CNS disease other than neoplasia, and the analysis of the B cell and T cell repertoire in those patients can provide meaningful information for diagnosis and management^53–58^. This is particularly relevant to autoimmune, inflammatory and infectious diseases affecting the CNS.

With these considerations in mind, we sought to establish an approach that could supplement neurosurgical biopsy by identifying and characterizing brain tumors through interrogation of genetic and immune cell profiles in CSF. There are several key capabilities that would be required for such an assay: 1) identification of driver mutations across a wide array of primary and metastatic cancers, 2) genome wide identification of chromosomal copy number alterations, 3) characterization of B and T cell populations, 4) robust compatibility with a relatively small amount of CSF DNA, 5) a uniform workflow that could use the same starting material for all analyses to minimize cost while maximizing ease, yield, sensitivity, and reproducibility, and 6) an informatics pipeline capable of analyzing and integrating these heterogenous datasets. The design and execution of such an approach is described in this manuscript (Fig. 1). We show that the assay developed here is able to identify brain tumors and provide clinically actionable information from CSF liquid biopsy rather than neurosurgical tissue biopsy.

**Figure 1.**
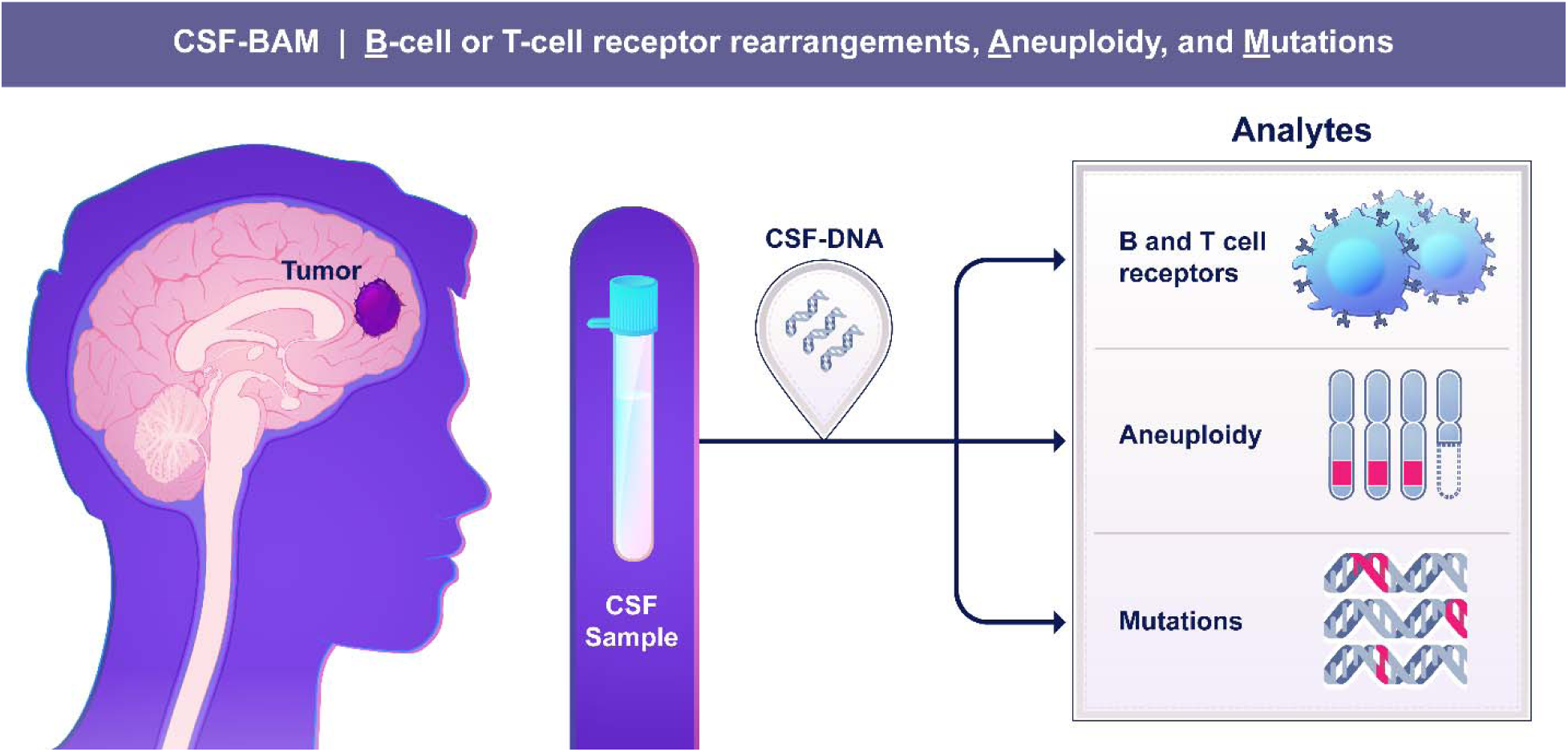
CSF-BAM overview. CSF is obtained and DNA is extracted from the entire sample. CSF-BAM examines three analytes simultaneously: **B** cell or T cell receptor rearrangements, **A**neuploidy, and **M**utations using PCR-mediated amplification of both strands of the DNA.

## Results

### Overview

Once DNA was purified from CSF, a DNA library was generated through a modified version of a protocol described previously, named SaferSeqS, which produces a relatively high conversion efficiency of the original DNA template molecules into library DNA molecules (see Methods)^59^. This conversion efficiency was particularly important when the quantity of CSF fluid was limited or when the DNA concentration in that fluid was low. Equally importantly, the SaferSeqS library preserved DNA from both the Watson and Crick strands of the original DNA templates. The ability to independently assess *both* strands of DNA exponentially increases the accuracy of the resulting sequencing data when the fraction of aberrant DNA molecules is low^59–61^. SaferSeqS libraries contain ∼200 copies of each of the original template molecule strands, and therefore can be used for multiple downstream assessments of DNA^59^. For CSF-BAM, we chose to analyze the clonal composition of DNA derived from malignant or normal B or T cells, as well as chromosome copy number alterations and somatic mutations derived from the cancer cells (Fig. 2). The paradigm for evaluation of each of these three components was identical and was performed in distinct stages to maximize reproducibility and minimize overfitting (Supplementary Table S1):

i. Analytical stage: optimize the experimental procedures and bioinformatic analysis using DNA from peripheral blood leukocytes or plasma cell-free DNA from healthy individuals.
ii. Training stage: use the optimized procedure to evaluate DNA from CSF or blood from a different cohort of patients with and without cancer to establish thresholds for specificity and estimate sensitivity at the chosen thresholds.
iii. Validation stage: use the optimized procedure to evaluate CSF from an independent cohort of patients to determine sensitivity and specificity at the pre-defined thresholds for positivity.

**Figure 2.**
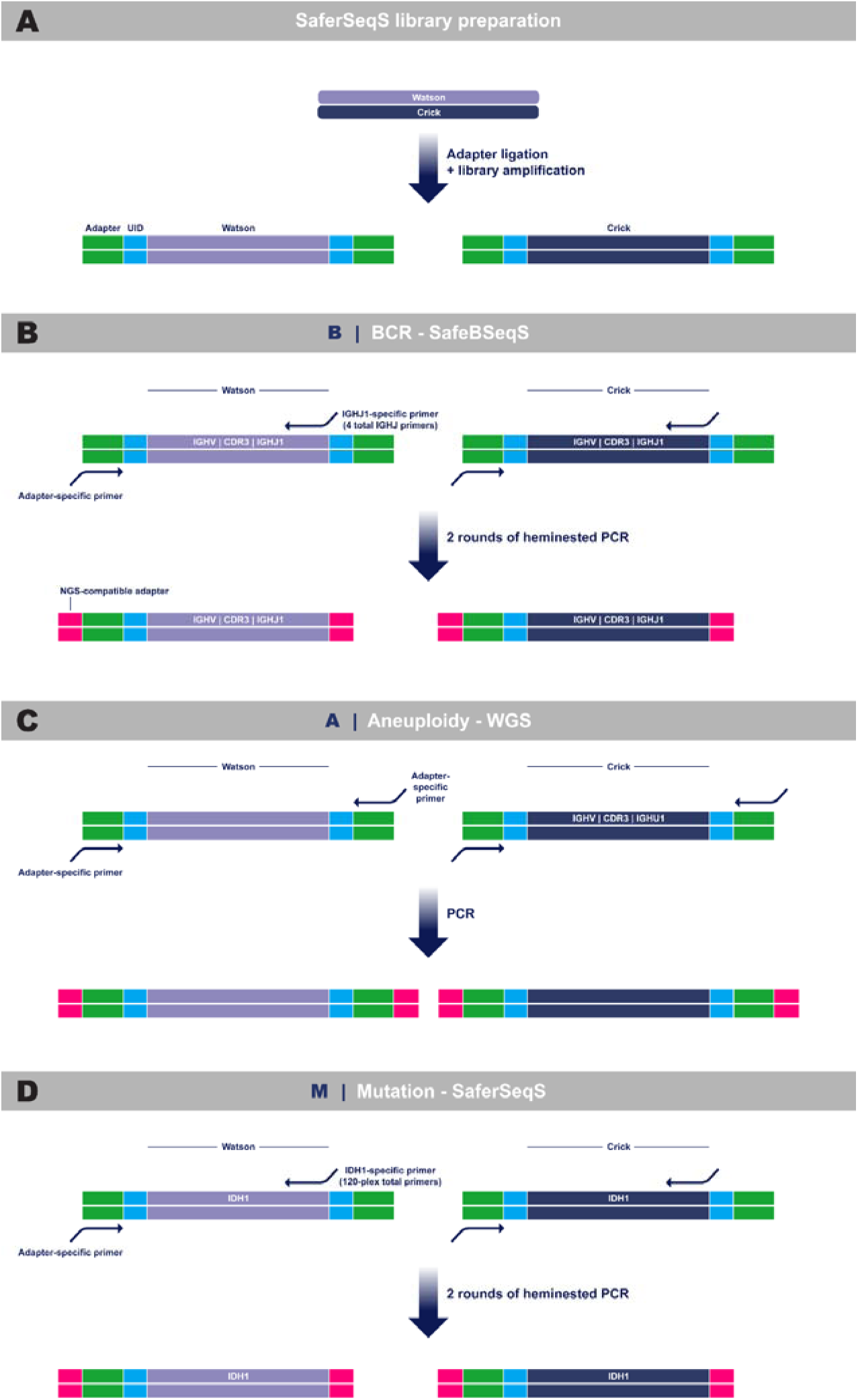
Schematic of CSF-BAM. **A,** Independent libraries are generated from each strand of the original DNA template molecules in a manner where unique molecular identifiers (UIDs) allow the strands from the two libraries to be mapped back to their original duplex. **B,** BCRs are evaluated with SafeBSeqS. Illustrated here is amplification of BCRs with a primer targeting the IGHJ1 segment, among the multiplex set of 4 total IGHJ primers. **C,** Aneuploidy is evaluated with WGS through amplification of total libraries using adapter-specific primers. **D,** Mutations are evaluated with SaferSeqS. Illustrated here is amplification of IDH1 using an IDH1-specific primer, among the multiplex set of 120 total gene-specific primers.

### Development of the experimental procedures and bioinformatic pipelines for CSF-BAM

The workflow for all three components of CSF-BAM begins with creation of libraries from original DNA template molecules (Fig. 2A). Copies of the DNA templates in the library, which are mappable to the original templates, are then split for analysis with each of the “B,” “A,” and “M” components.

#### The “B” component of CSF-BAM

queries the B cell (BCR) and T cell (TCR) receptor genes that are integral to the adaptive immune system. In any single individual, millions of BCR and TCR receptors in normal B and T cells, respectively, are generated through imprecise joining of variable (V), diversity (D), and joining (J) segments of the BCR or TCR genes. The nature of these sequences and the degree of clonality provides a wealth of information about the adaptive immune system in that particular patient. In comparison to flow cytometry, TCR or BCR sequencing provides comprehensive repertoire descriptions^62^. Moreover, because any B cell or T cell cancer is derived from a single B cell or T cell, respectively, a neoplastic clone is characterized by a single VDJ rearrangement. The presence of malignant cells in CSF can thereby be detected by the over-representation of a single sequence in the CSF, implying a predominant clone. Although such clones can be detected through previously published methods that sequence either RNA or DNA templates, accurate detection and quantification of clonotypes is challenging. Among the reasons for this is that sequencing from DNA templates has generally required multiplex combinations of primers to amplify all possible V and J gene segment pairs^63–65^. SafeBSeqS and SafeTSeqS overcome this challenge by requiring primers for only the J segments of one of the BCR (*IGH*) or one of the TCR (*TCRB*) genes, respectively.

Though these data cannot be used to analyze the constant region, they can be used to re-construct the entire VDJ sequence of the TCR or BCR. They are therefore adequate to determine the clonal representation of any population of B cells or T cells as well as to identify certain characteristics of the rearrangements associated with cancers or autoimmune disease ^66^.

After extensive experimentation in the analytical stage, we found that four primers were sufficient to assess the entire BCR repertoire with SafeBSeqS (Supplementary Table S2A). One primer amplified gene segments IGHJ1, IGHJ4, and IGHJ5, and one primer each amplified IGHJ2, IGH3, and IGHJ6. These four primers were mixed together and used for hemi-nested amplification of the SaferSeqS libraries. They yielded uniform amplification of all the queried gene segments as tested on a sample of DNA derived from fibroblasts with a uniform representation of gene segments (Supplementary Fig. S1). For the training stage, we then applied SafeBSeqS to the evaluation of DNA from peripheral white blood cells of 95 healthy control individuals and CSF from 25 individuals with primary and secondary CNS lymphoma as a training set. The summaries of results are listed in Supplementary Table S3 and Supplementary Table S4. Based on these results, we defined the positive criteria for clonality as total UIDs ≥20 and top clone UIDs / total UIDS ≥0.3 (30%). In total, 1/95 samples from healthy individuals and 13/25 samples from patients with CNS lymphoma in our set met these criteria (Supplementary Tables S3 and Supplementary Table S4). For the validation stage, when SafeBSeqS was applied to CSF from a different cohort of individuals without known cancers, we found that 0 met the positive criteria for clonality (Supplementary Table S4). Similarly, 1 of the 202 CSF samples from patients with cancers other than CNS lymphomas scored positively (Supplementary Table S4). Of 4 CSF samples from patients with B cell lymphomas of the CNS, 2 scored positively in this assay with clonal fractions of 32% and 86% (Supplementary Table S4).

In the analytical stage for SafeTSeqS, we found that 13 primers, one for each TRBJ gene segment, were sufficient to assess the TCR repertoire (Supplementary Table S2B). Uniform amplification of all 13 J segments was obtained in control fibroblast DNA (Supplementary Fig. S2). Supplementary Table S5 lists the summary data of SafeTSeqS data obtained from the evaluation in the training stage of DNA from peripheral white blood cells of 95 healthy control individuals. Using the same criteria as for SafeBSeqS, we defined positive clonality as total UIDS ≥20 and top clone UIDs / total UIDs ≥0.3 (30%). Zero samples met these criteria. When SafeTSeqS was applied for the validation stage to CSF from a different cohort of 33 samples from individuals without known cancers, we found that 0 samples met the clonality criteria (Supplementary Table S6). Three of the 202 CSF samples from patients with cancers other than CNS lymphomas scored positively in this assay (Supplementary Table S6). No patients with T cell lymphomas of the CNS were available to us, so we assessed peripheral white blood cell DNA from 3 patients with peripheral T cell neoplasms who had clonal T cells present in the blood on clinical flow cytometry testing. Of 3 such patients, 3 scored positively in this assay, with clonal fractions of 55%, 57% and 95% (Supplementary Table S5).

#### The “A” component of CSF-BAM

queries aneuploidy throughout the genome. SaferSeqS libraries are converted to a form suitable for whole genome sequencing (WGS) by the addition of primers whose sequences match those of the Illumina NovaSeq flow cells (Fig. 2C and Supplementary Table S2C). The WGS data are mapped to the human genome through standard methods. A modified version of the ichorCNA algorithm^67^ is used to assess gains or losses on 34 chromosome arms. Sex chromosomes, acrocentric chromosomes, and arms with high background are excluded in the analysis (see Methods). Though Watson and Crick strands can be identified from the sequencing data, there is no need to couple the reads to ensure single base pair accuracy to assess copy number alterations of entire arms.

For the training stage we applied this WGS assay to SaferSeqS libraries from CSF of 31 individuals without cancer (training cohort—Supplementary Table S7). We used two metrics of aneuploidy to derive a threshold for scoring samples as aneuploid. First, we evaluated the number of arms altered. In our training cohort, two of the 31 samples had exactly one arm altered while the remaining 29 samples had no arms altered. As a result, any sample with more than one arm altered would be scored as positive for aneuploidy. Next, we evaluated the estimated tumor fraction as predicted by ichorCNA. In our training cohort of individuals without cancer, the largest predicted tumor fraction was 1.4%. Any sample predicted to have a tumor fraction >1.5% would be scored as positive.

When the same assay was applied for the validation stage to WGS data derived from SaferSeqS libraries of CSF from a different cohort of individuals without active cancers, we found that 0 of 33 scored positively (100% specificity, credible interval 90% to 100%) (Supplementary Table S8). 121 of 205 CSF samples from patients with CNS cancers of various types had detectable aneuploidy. Of these, 55 of 81 samples from patients with high grade gliomas were positive (68%, credible interval 57% to 77%). Patients with medulloblastomas or cancers outside the CNS that had metastasized to the brain also were often positive for aneuploidy using this measure (81%, credible interval 60% to 92% and 85%, credible interval 66% to 94%, respectively, Supplementary Table S8).

The reproducibility of the approach was assessed through the evaluation of independent aliquots of CSF DNA as technical replicates from the same patient (n = 104). Each aliquot had an independent SaferSeqS library. For the non-cancer controls (n=25) with a technical replicate, all arms were concordant while 98.5% of the arms in the cancer technical replicates (n=79) were concordant (Supplementary Table S9; Supplementary Fig. S3; Methods). A subset of samples (n=30) were previously described and evaluated using the Repetitive Element Aneuploidy Sequencing System (RealSeqS)^32^. RealSeqS uses a single PCR primer to concomitantly amplify ∼350,000 loci spread through genome in order to evaluate aneuploidy^68^. We compared the chromosome arm level calls between the two assays and found 94.1% of the arms were concordant (Supplementary Table S10; Methods). Of the discordant calls, a majority could be explained as falling just below the threshold for one of the two assays.

#### The “M” component of CSF-BAM

identifies subtle somatic mutations such as single base substitutions (SBS) or small insertions or deletions (indels). For this component, extremely high specificity is required to minimize errors during the experimental or bioinformatic components of this assay. The workflow is identical in principle to that described above for SafeTSeqS and SafeBSeqS (Fig. 2B and D) but different primers are used. Instead of 4 primers for SafeBSeqS and 13 primers for SafeTSeqS, 120 primers were used for mutation analysis, each amplifying a region of the genome that is commonly mutated in cancers of the CNS or cancers that metastasize to the CNS (Fig. 2D). These primers (Supplementary Table S2D) were chosen after extensive experiments in the analytical stage to maximize the uniformity of representation of the amplicons as well as minimize the number of off-target reads upon sequencing. The uniformity of amplification of the 120 amplicons queried by the 120-plex is shown in Supplementary Fig. S4.

It is well known that mutations in cell-free DNA (cfDNA) from peripheral blood largely arise from either tumors or CHIP (Clonal Hematopoiesis of Indeterminate Potential)^69^. To help ensure mutations identified in the CSF were not a result of CHIP, we analyzed matched WBC DNA in available cases as previously reported^70^.

For the training stage we evaluated CSF from a different cohort of 300 individuals without cancer (Supplementary Table S11). These samples do not have matched peripheral blood to eliminate CHIP mutations and were not used for the evaluation of performance metrics. We used this cohort to tune our somatic mutation calling approach and determine thresholds for positivity (training set). We report 20 mutations in *TP53*, 3 in *KRAS*, 5 *NRAS*, 8 mutations in *FBXW7* in codon 505, and 7 in other amplicons. Given the abundance of non-canonical mutations in KRAS and NRAS in the non-cancers, we restricted future mutation calls within these genes to only KRAS codon 12 and NRAS codon 61, which represent the most commonly mutated hotspots in cancer.

The FBXW7 codon 505 mutations in the non-cancer samples were unexpected. This specific codon is not typically mutated in CHIP and other codons throughout FBXW7 are typically mutated in cancer. Upon closer inspection, every molecule with a mutant FBXW7 codon 505 also had mutations at codons 289, 299, 300, and 314. We performed blat analysis to all possible non-human genomes^71^. The sequence from the observed mutated FBXW7 molecules perfectly matched the bovine genome. We hypothesized that bovine-derived hemostatic agents frequently used in neurosurgery^72^, when CSF was collected for the trigeminal neuralgia samples, may have contributed minor amounts of DNA that amplified and incorrectly aligned to FBXW7.

On the basis of these data, we chose to positively score mutations that were present in more than one original template molecule and found in cancer patients in the COSMIC database (see Methods). Given the importance of TERT promoter mutations in CNS neoplasms, we relaxed this metric to score samples with even one mutant template molecule as positive.

Using these thresholds for the validation stage, we identified somatic mutations in 0 of 33 individuals without cancer (credible interval 90% to 100%) (Supplementary Table S12). 79 of 206 CSF samples from patients with CNS cancers of various types had detectable mutations (38%, credible interval 32% to 45%). Of these, 44 of 81 samples from patients with high grade gliomas were positive (54%, credible interval 43% to 65%). Of the 5 high grade gliomas that were scored positive based on the presence of only one mutant TERT molecule, all had the canonical gain on chr7 but 4 of 5 fell just below the threshold for aneuploid positivity. Given the heterogenous nature of medulloblastoma driver mutations, only 4 of 21 (19%, credible interval 8% to 40%) samples scored positive for mutations. 17 of 24 metastatic cancer samples (71%, credible interval 51% to 85%) had mutations detected.

### Application of CSF-BAM to CSF samples (validation stage)

We evaluated 239 CSF samples from 222 patients; for each sample, peripheral blood WBC DNA was available to exclude any mutations due to CHIP. Clinical information including demographics are described in Supplementary Table S13, Supplementary Table S14, and Supplementary Table S15. The amount of CSF available for these studies averaged 3.5 mL and ranged from 0.5 to 14 mL. The amount of DNA recovered from CSF averaged 25 ng (IQR 2.6 to 29.6 ng) (Supplementary Table S15). Supplementary Table S15. also includes summaries of the sequencing data obtained from all patients, and whether they scored positively in the B, A, or M components of CSF-BAM. If a patient scored positively in at least one of these assays using the pre-defined thresholds described above, the patient was considered positive for CSF-BAM. Of note, 82% (119/146) of samples derived from individuals with brain cancers, defined as grade 3 or 4 for primary brain tumors, were positive.

#### 33 samples from patients without cancer

None of the CSF samples from these patients scored positively in any of the three components, yielding 100% specificity (credible interval 90 to 100%) (Fig. 3 and 4 and Supplementary Table S15).

**Figure 3.**
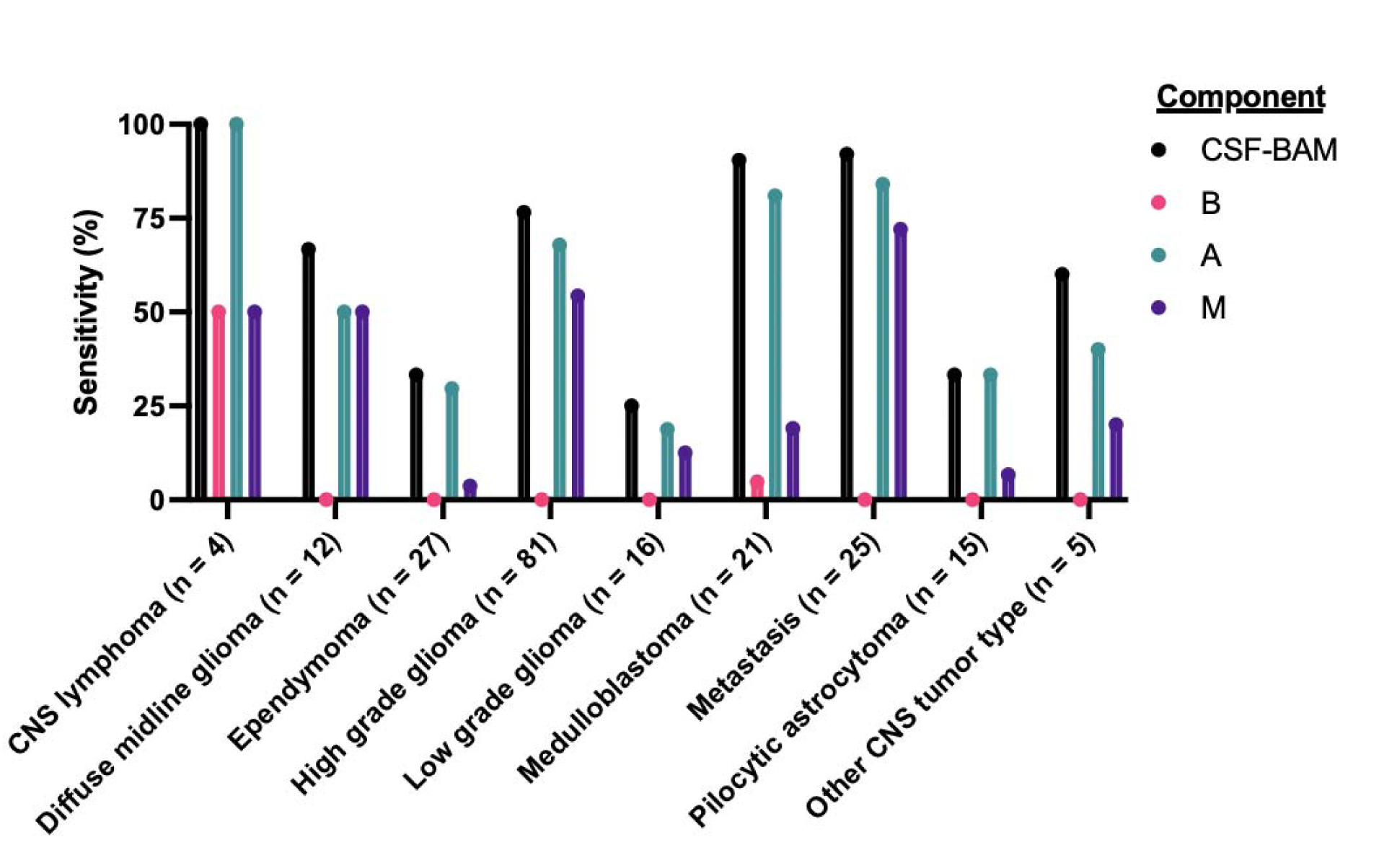
Performance of CSF-BAM and each analyte. The sensitivity of each analyte within CSF-BAM is demonstrated across the major class of tumors tested. The composite sensitivity of CSF-BAM is demonstrated in black.

**Figure 4.**
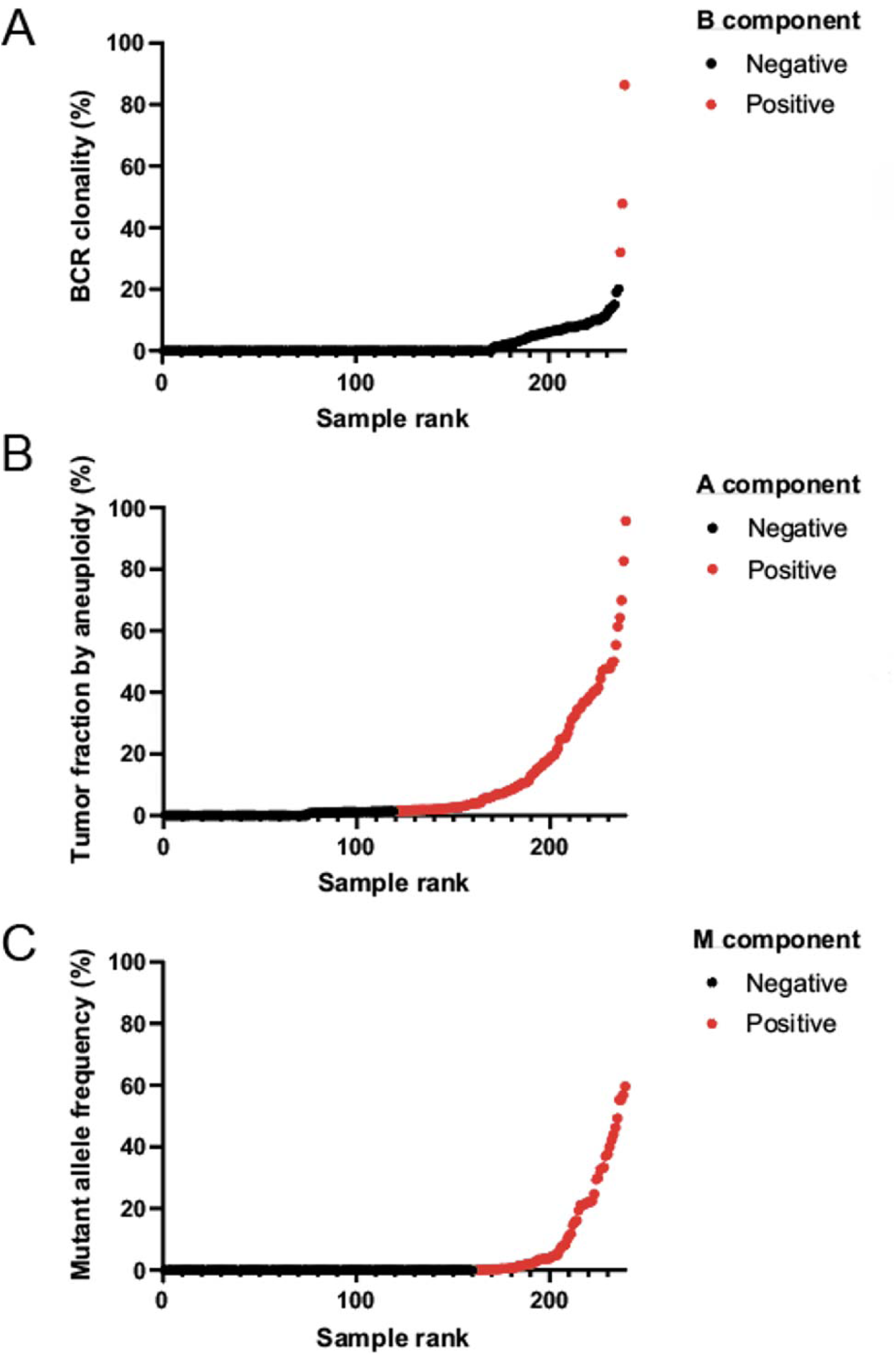
**A,** BCR clonality (clonality for non-evaluable samples with total UIDs ≤20 defined as 0), **B,** estimated tumor fraction, and **C,** mutant allele frequency for each sample evaluated with CSF-BAM.

#### 81 samples from patients with high grade glioma

62 of these samples scored positively for at least one of the three components, yielding a sensitivity of 77% (credible interval 66% to 84%) (Fig. 3 and 4 and Supplementary Table S15). Notably, 82% (54/67) of glioblastomas were detected via CSF-BAM. No sample scored positively with the B component, as expected. The aneuploidy component was the predominant basis for sensitivity, with 67% positive (credible interval 56% to 76%). The mutation component scored positively in 44 samples, including 7 that were not scored positively by aneuploidy.

#### 21 samples from patients with medulloblastomas

19 of these samples scored positively for at least one of the three components, yielding a sensitivity of 90% (credible interval 71% to 97%) (Fig. 3 and 4 and Supplementary Table S15). One sample scored positively with the B component. The aneuploidy component was the predominant basis for sensitivity, with 81% positive (credible interval 60% to 92%). The mutation component scored positively in 4 samples, including 2 that were not scored positively by aneuploidy.

#### 25 samples from patients with metastatic lesions to the brain

23 of these samples scored positively for at least one of the three components, yielding a sensitivity of 92% (credible interval 75% to 98%) (Fig. 3 and 4 and Supplementary Table S15). No sample scored positively with the B component, as expected. The aneuploidy component was the predominant basis for sensitivity, with 54% positive (credible interval 65% to 93%). The mutation component scored positively in 17 samples, including 1 that was not scored positively by aneuploidy.

#### 79 samples from patients with other tumor types

These patients included those with CNS lymphomas, gliomas other than high grade, ependymomas, and various other primary brain tumor types (Supplementary Table S15). 33 of these samples scored positively for at least one of the three components, yielding a sensitivity of 42% (credible interval 32% to 53%) (Fig. 3 and 4 and Supplementary Table S15). As with high grade gliomas, medulloblastomas, and metastatic cancers, the aneuploidy component was the predominant basis for sensitivity, with 35% positive (credible interval 25% to 35%). The mutation component scored positively in 13 patients, including 5 that were not scored positively by aneuploidy.

#### Other genetic observations of interest

When detectable by the A component, the median neoplastic DNA fraction based on the analysis of aneuploidy was 8.2% (IQR 2.5% to 26.9%). When detectable by the M component, the median mutant allele fraction based on the analysis of mutations was 3.9% (IQR 0.7% to 21.9%). The correlation between the two genetically altered fractions was high (R=0.55, P<5e-6; Supplementary Fig. S5).

The nature of the genetic alterations provided some insight into the type of tumor present in the CNS. Mutations in IDH1 at codon 132 or 172 were observed in 8 samples, and all (100%) of these were from patients with gliomas (2 with oligodendroglioma WHO Grade 3, 1 with astrocytoma WHO grade 3, and 5 with astrocytoma WHO grade 4). In all of these cases, standard sequencing of the resected gliomas revealed IDH mutations. In 3 additional samples from individuals with IDH mutant gliomas, no IDH mutations were observed in the CSF. Histone H3F3A mutations at codon 28 were detected in the CSF of 6 patients (3 with diffuse midline gliomas, 3 with GBM). Two of the six patients had their tumor sequenced, and in both cases the identical mutation was found in the matching CSF. However, no H3F3A mutations in the CSF were described in 5 other individuals who were diagnosed with H3.3 mutated gliomas. KRAS mutations at codon 12 were noted in 3 patients which harbored metastatic lesions from outside the CNS.

### Relationship of CSF-BAM results to clinical characteristics

Cytology is not routinely performed on all CSF at our institutions given its poor performance but was available in 53 cases, 50 of which were from individuals with cancer (Supplementary Table S15). The three cases without cancer were negative by cytology as well as by the CSF-BAM assay. Cytology was positive in nine (19%) of the cancer cases, and eight of (89%) were also scored as positive by the CSF-BAM assay. Of five cases recorded as “suspicious” on cytology, four (80%) were scored as positive by the CSF-BAM assay. Of 36 cases with cancers diagnosed as negative by cytology, 19 (53%) were scored as positive by CSF-BAM. Overall, CSF-BAM was positive in 24 cancer cases where cytology was negative or inconclusive, while only missing one cancer case of 9 that were positive by cytology.

We additionally evaluated whether tumor contact with the CSF space was related to positivity by CSF-BAM (Supplementary Table S16). We compared all samples from patients with cancer for which clinical data was available. Using MRI contrast enhancement as a tumor marker, tumors abutting the CSF space (cistern, ventricle or cortical surface) were more likely than those not abutting the CSF space to be positive by CSF-BAM (P = 0.006 by Fisher’s exact test, odds ratio = 4.1, 95% confidence interval 1.5-10.5). Similarly, using MRI T2 signal as a tumor marker, tumors juxtaposed to a CSF space were also more likely to be positive by CSF-BAM (P = 0.03 by Fisher’s exact test, odds ratio = 4.8, 95% confidence interval 1.3-16.9).

### Immune receptor repertoire profiling

We evaluated a total of 264 CSF samples using SafeBSeqS for the B component of CSF-BAM in addition to SafeTSeqS for TCRs (Supplementary Tables S4, S6, and S15). We identified a mean of 289 (range 0-3701) total UIDs per sample representing original TCR DNA template molecules and a mean of 36 (range 0-1787) total UIDs per sample representing original BCR DNA template molecules (Supplementary Fig. S6). Because each unique UID can only be derived from a single cell in a SaferSeqS library ^59^, these data can be used to determine the number of mature B or T cells present in the CSF when accounting for input amounts of CSF. Few samples had high proportions of distinct clones of T-cells (Supplementary Fig. S7A). However, the sequencing data revealed 187 TCR specificity groups, composed of at least 3 TCRs from patients with cancer, as determined by TCR amino acid sequence motifs, suggesting shared activity (Supplementary Table S17A)^73^. The top TCR clone in only 2 samples had a match with high or very high confidence for sequences of previously identified TCRs with annotated specificity (Supplementary Table S17B)^74^.

B cell clonality varied substantially by cancer type (P < 0.001 by Kruskal-Wallis test) (Supplementary Fig. S7B). Using clonality as a metric to classify CNS lymphomas against all other cancer types for evaluable samples produced a ROC curve with an AUC of 0.98 (95% confidence interval 0.93-1.00) (Supplementary Fig. S8A). The proportion of samples for which the most frequently observed clone contained the IGHV4-34 gene segment was significantly enriched in CNS lymphomas compared to all other sample types (P = 0.0029 by Fisher’s exact test) (Supplementary Fig. S8B). This degree of IGHV4-34 gene segment representation was similar to that observed in prior studies of both peripheral and CNS lymphomas^75,76^.

### Case reports demonstrating clinical applicability

We highlight three cases within this cohort that demonstrate the potential clinical applicability of CSF-BAM. GLIA793 is a sample from a patient with grade 3 oligodendroglioma who underwent previous resection followed by adjuvant radiation and chemotherapy. CSF-BAM was negative on CSF obtained three weeks prior to repeat resection when there was a concern for tumor progression (Supplementary Fig. 9A). Pathological examination of the surgical specimen revealed treatment effect without evidence of recurrent disease, a common finding in post-treatment gliomas^77^. Another example is GLIA 914, an individual with metastatic breast cancer to the brain with suspicion of leptomeningeal disease (LMD). The patient had over five lumbar punctures for evaluation of LMD via cytology over a 12-month period and each was negative. The final CSF sample was tested via cytology and also CSF-BAM. The cytology was indeterminant with only rare atypical cells. However, this sample was robustly positive via CSF-BAM with a tumor fraction estimated to be approximately 40% with both aneuploidy and mutations detected. A third case is GLIA 886, derived from an individual with a spinal cord ganglioglioma. The patient had a slowly recurrent spinal cord tumor which was initially resected many years ago prior to the advent of routine tumor sequencing and treated with carboplatin. The tumor was not responsive to chemotherapy and the individual underwent a repeat resection which demonstrated a BRAF mutant ganglioglioma (Supplementary Fig. 9B). CSF-BAM identified the same disease defining canonical BRAF p.V600E mutation in CSF obtained at the time of surgery.

## Discussion

Minimally invasive methods to detect brain cancers is a major unmet need in neurology and neuro-oncology. The current standard of care requires invasive neurosurgical biopsy with associated risk and cost. Our group and others have previously demonstrated that CSF is enriched for tumor associated genetic and epigenetic alterations^32,33,78–80^. Other studies have shown that the evaluation of B cells and T cells in CSF can provide valuable information about the pathogenesis of neoplastic and non-neoplastic diseases^52,53,81^.

We here describe a unified approach that can comprehensively assess CSF DNA for somatic mutations and aneuploidy as well as enumerate clonal compositions of both B cells and T cells. The workflow incorporates several novel features and can be applied to relatively small amounts of CSF. The entire assay can easily be performed within a week of obtaining CSF because a single DNA library, including amplification of both strands of the entire genome, is used for all its component assays.

The clinical scenarios in which CSF-BAM could be applied are vast. The results of our study reveal the multiple ways in which CSF-BAM could impact clinical care. One way is by providing potential for diagnosis of brain cancers before tissue biopsy, with information added beyond what can be obtained from imaging. For the 82% of samples from patients with cancer that were positive by CSF-BAM in our study, neoplasia could have potentially been diagnosed based on CSF sampling and imaging prior to neurosurgical biopsy. In important situations patients could have been spared from tissue biopsy, as illustrated for GLIA793. In this case, CSF-BAM could have provided evidence for pseudoprogression despite increasing contrast enhancement on MRI scan. Pseudoprogression is a common challenge in both primary and metastatic brain tumors treated with chemotherapy and/or radiation therapy. Having additional molecular tools to better identify pseudoprogression without tissue sampling would be clinically meaningful.

A second way in which CSF-BAM could improve upon current clinical care is by providing more sensitive and detailed diagnoses than can be obtained through cytology. Although cytology is often used to assess CSF samples for the presence of malignant cells and is highly specific, it requires large volumes of CSF (>8 mL)^82^, its sensitivity is relatively low, and results of uncertain significance (“suspicious for malignancy”) are often reported. Flow cytometry can add diagnostic value in some settings^35^. In the current study, we show that most (25 of 33) cases of CNS cancers that were detected with CSF-BAM were *not* detected by cytology. Conversely, CSF-BAM detected the great majority (8 of 9) cancers positive by cytology, and was positive in 4 of 5 cases which were classified as “suspicious” by cytology. Though the sensitivity of CSF-BAM is considerably higher than that of cytology, it would be prudent to include both types of assessments for diagnosis in future studies. The potential utility of CSF-BAM is illustrated by the case of GLIA 914, a patient with leptomeningeal metastasis who was positive with CSF-BAM. Prognoses for leptomeningeal metastases are extremely poor in part due to the difficulties in diagnosis. Indeed, this patient had multiple negative cytologic examination over many months. CSF-BAM in this instance could have provided swifter diagnosis. Earlier detection can enable more rapid initiation of treatment to preserve neurological function and potentially improve survival.

A third impact from CSF-BAM is from the detailed genetic characterization provided by the assay. The molecular composition of CNS neoplasms can provide opportunities for rational and biologically guided treatments ^83^. For example, targeted therapies are showing increasing success in neuro-oncology ^90^. Having an assay like CSF-BAM to diagnose the BRAF mutation in GLIA 886 could have enabled treatment with BRAF inhibitors, which have shown promise in gangliogliomas and if successful could have delayed or obviated the need for a repeat resection^84^. In addition, neurosurgical biopsies are often small and don’t provide definitive diagnosis as in the case of GLIA 58, an adult with a spinal cord tumor. The patient underwent surgical biopsy but the pathological analysis returned as glial hypercellularity with features of a diffuse astrocytoma, an equivocal finding. CSF-BAM was positive and if combined with tumor tissue testing could have enabled a definitive diagnosis.

The sensitivity of CSF-BAM was high for the major cancer types that would prompt evaluation of CSF during the diagnostic work-up. In particular, more than 80% of GBM, medulloblastomas, CNS lymphomas, and metastases from primary tumors outside the CNS could be detected (Fig. 3 and Supplementary Table S15). As ours was a retrospective study based on available samples rather than a prospective study, the fraction of the various tumor types evaluated was different than what would be seen in practice. If corrected for incidence according to the Surveillance, Epidemiology and End Results (SEER) Database, we would have about 10-15 times the number of metastatic cases compared to the primary CNS cancers, which would impact overall test performance metrics given that brain metastases were correctly detected in over 90% of tested samples.

The limits of sensitivity for detecting malignancy in the CSF are not yet known. From studies of pancreatic, colorectal, breast, and other cancer types in plasma, it is known that some advanced cancers do not release (“shed”) DNA into the plasma^85^. Analogously, some CNS cancers may not release DNA into the CSF, especially those that are not anatomically adjacent to a CSF space^33^. Improvements in CSF-BAM could increase sensitivity further. One simple improvement would be the analysis of a higher volume of CSF; many of the samples in our retrospective collection included <1 mL of CSF, which limits the number of molecules that can be assessed, particularly for mutations or B or T cell clonality. In addition, applying metrics for minimum input DNA would help ensure sufficient starting material is available to reliably detect the three BAM analytes. Another improvement would be the inclusion of additional clinically relevant oncogenes and tumor suppressor genes in the mutational analysis.

A third potential improvement could be the inclusion of epigenetic alterations into the analysis. This would technically be straightforward, as SaferSeqS library preparation can be easily modified to permit evaluation of DNA methylation as well as of aneuploidy and mutations^86^. DNA methylation could also help determine the CNS tumor type present, analogous to plasma cfDNA used to determine the tumor of origin of non-CNS cancers^87,88^. A caveat to using methylation is the high analyte sensitivity required to detect cases. In our study, most of the cancers that were detected by CSF-BAM were detected through aneuploidy rather than mutations or immune cell clonality. To be scored as positive for aneuploidy generally required a 1.5% tumor fraction. In other words, if < 1.5% of the DNA in the CSF did not arise from cancer aneuploid cells, then it would generally not be scored as positive. 15 cancers not detected by the aneuploidy component were detected by mutations (Fig. 3 and 4 and Supplementary Table S15) and the limit of detection for mutation was as low as 0.01% in this cohort. If methylation is to meaningfully add sensitivity to CSF-BAM, the methylation assay would have to be capable of detecting differentially-methylated regions of the genome present in <1% of the template molecules, while retaining a specificity close to 100%, which is challenging.

Though there is strong and increasing interest in sequencing of TCRs and BCRs to understand the immune components of disease, current technologies to sequence immune receptors suffer from limitations in throughput, sensitivity, reproducibility, and abundance quantification^63,64^. Previous studies of immune receptor repertoires in CSF have generally focused on aging, neurodegeneration, and neuroinflammatory conditions^57,58,89^. In patients with cancer, studies have primarily reported TCR and BCR sequences derived from cells in tumor tissue with relatively limited data reported on immune receptor repertories derived from CSF^81,90–94^. We substantially add to this information through the analysis of BCRs and TCRs in 231 CSF samples from patients with neoplasms. SafeBSeqS and SafeTSeqS, the new technologies described in this manuscript, use DNA rather than RNA as the analyte. Combined with the high conversion of template molecules to library molecules and the sequencing of both strands of DNA to limit errors made during library preparation and sequencing, these tools provide a way to quantify the number of B cells or T cells in any bodily compartment, as well as the repertoire of the TCRs and BCRs encoded by those cells. The number of B cells or T cells in a sample cannot be reliably assessed with RNA-based technologies because the amount of RNA can vary substantially among cells, especially in complex environments such as the CNS. In contrast, each mature B cell or T cell generally contains a single rearranged BCR or TCR gene, so the number and fraction of UIDs in the sequencing data provides a direct estimate of the number of B cells and T cells in the original sample.

Limitations of our study include the relatively small sample size for several tumor types. It is conceivable that the pre-operative results of CSF-BAM, and its degree of positivity, provide information about prognosis for some cancer types, but our study was not powered to do so. Moreover, we studied only a single sample of CSF for most patients. Serial, longitudinal studies of patients post-treatment would be required to determine whether the analysis of CSF will provide information about the effectiveness of treatment on patients with CNS tumors, which is often challenging based on radiographic findings alone. Finally, CSF samples from many more patients without cancer will be required to obtain a more precise estimate of specificity than the one we currently have, i.e., 100%, but with a lower bound of the credible interval at 90%.

One of the strengths of our study was the segregation of available samples into independent analytical, training, and validation cohorts, permitting pre-defined thresholds to be applied to the validation cohort^95^. Another strength is that the libraries used to implement CSF-BAM provide a limitless source of DNA from the CSF, given that there is a 1:1 correspondence between both the Watson and Crick strands of the template DNA molecules and the molecules in the amplified library. PCR-mediated re-amplification of the original libraries would thereby permit analysis of the same precious CSF samples with improved versions of CSF-BAM, as well as permit sharing of samples among laboratories. For these reasons, as well as its encouraging performance on CSF samples to date, CSF-BAM provides a promising foundation for the molecular genetic evaluation of the DNA from CSF samples. The potential for CSF-BAM integration into clinical workflows will be the subject of future prospective studies.

## Methods

### Study design

This study was approved by the Institutional Review Boards for Human Research at Johns Hopkins Hospital (IRB00075499, NA_00090530, and IRB00292573) in compliance with the principles of the Declaration of Helsinki and the Health Insurance Portability and Accountability Act. All patients provided written informed consent. All samples were deidentified immediately following collection. All available samples were included in the study. Clinical data for the individuals included in the study are presented in Supplementary Table S10. Some samples have been previously studied in prior publications using different technologies^32,33^. Importantly, all samples in this manuscript had new CSF-BAM libraries generated as described below from CSF derived DNA.

### Sample processing and DNA purification

CSF samples were collected into standard CSF collection tubes. Volumes used for DNA purification are reported in Supplementary Table S15. Blood samples were collected into Streck Cell-Free DNA BCT (#230469). DNA from CSF, plasma, or leukocytes was purified using the BioChain Cell-free DNA Extraction Kit (#K5011625). Control primary dermal fibroblasts were obtained from ATCC (#PCS-201-012) and DNA was purified using Qiagen QIAamp DNA Mini Kit (#51304).

### Library construction

We developed a library preparation workflow that can efficiently recover input DNA and simultaneously incorporate double-stranded molecular barcodes^59^. In brief, libraries were prepared using an Accel-NGS 2S DNA Library Kit (Swift Biosciences, 21024) with the following critical modifications: 1) DNA was pretreated with 3 U of USER enzyme (New England BioLabs, M5505L) for 15 min at 37 °C to excise uracil bases; 2) the SPRI bead/PEG NaCl ratios used after each reaction were 2.0×, 1.8×, 1.2× and 1.05× for end repair 1, end repair 2, ligation 1 and ligation 2, respectively; 3) a custom 50 µM 3′ adapter was substituted for reagent Y2 and 4) a custom 42 µM 5′ adapter was substituted for reagent B2. Libraries were subsequently PCR amplified in 50-µl reactions using primers targeting the ligated adapters. The following reaction conditions were used: 1× NEBNext Ultra II Q5 Master Mix (New England BioLabs, M0544L), 2 µM universal forward primer and 2 µM universal reverse primer. Libraries were amplified with 8 or 11 cycles of PCR, depending on how many experiments were planned, according to the following protocol: 98□°C for 30□s, cycles of 98□°C for 10□s, 65□°C for 75□s and hold at 4□°C. If eight cycles were used, the libraries were amplified in single 100-µl reactions. If 11 cycles were used, the libraries were divided into eight aliquots and amplified in eight 50-µl reactions, each supplemented with an additional 0.5□U of Q5 Hot Start High-Fidelity DNA Polymerase (New England BioLabs, M0493L), 1□µl of 10□mM dNTPs (New England BioLabs, N0447L) and 0.4□µl of 25□mM MgCl2 solution (New England BioLabs, B9021S). The products were purified with 1.8× SPRI beads (Beckman Coulter, B23317) and eluted in EB buffer (Qiagen).

### The “B” component of CSF-BAM (BCR)

TCR and BCR sequences were amplified from libraries using gene-specific primers for TRBJ and IGHJ segments as previously described for sequencing of somatic mutations^59,86^. A total of 13 unique primers were used to cover all TRBJ gene segments in each amplification step and 4 unique primers were used to cover all IGHJ gene segments in each amplification step (Supplementary Table S2A and S2B). For clonotype analysis, demultiplexed reads were used to generate clonotype tables using the MiXCR 4.6.0 package^96^ with the function “analyze generic-amplicon-with-umi” and specifications “--species hsa --dna --rigid-left-alignment-boundary --floating-right-alignment-boundary J.” The specification “--tag-pattern ^(R1:*)\^(UMI:N{14})(R2:*)” was used for Watson strands and the specification “--tag-pattern “^(UMI:N{14})(R1:*)\^(R2:*)” was used for Crick strands. Clones with duplex support were assembled using the function “exportClonesOverlap” with the specifications “--criteria CDR3|NT|V|J” for each Watson and Crick sample pair. Total UIDs are reported as the sum of UIDs from Watson and Crick samples for clonotypes with duplex support. Clustering was performed using GLIPH2^73^ for productive TCR sequences from CSF samples from patients with cancer. Comparison of TCR sequences to a database of TCRs with known specificities was performed using VDJdb^74^. To determine the relative yield for IGHJ and TRBJ gene segments, we evaluated DNA from primary fibroblasts. Non-rearranged IGHJ and TRBJ gene segments were analyzed using the computational pipeline as described for the “M” component below, with the modification of mapping to hg38. IGHJ1, IGHJ4, and IGHJ5 were computationally distinguished using primer sequence inputs GAGGAGACGGTGACCAGGGTGCCCTGGCCCCAGTG, GAGGAGACGGTGACCAGGGTTCCCTGGCCCCAGTA, and GAGGAGACGGTGACCAGGGTTCCCTGGCCCCAGGG respectively in the analysis pipeline.

### The “A” component of CSF-BAM (aneuploidy)

Library DNA was amplified in 50 μL reactions in Ultra Q5 with primers at 2 μM for seven cycles with the following conditions: 98 °C for 30 s, then seven cycles of 98 °C for 10 s to denature, and 65 °C for 75 s to anneal and extend. WGS libraries were sequenced on a NovaSeq 6000 with paired-end 2×100bpreads. The depth of sequencing averaged 40.0 M reads of 100 bp (IQR 33.8M to 45.1M, Supplementary Table S8). Cutadapt was used to trim 27 base pairs from both reads ^97^ and BWA-MEM was used to align reads to the hg19 genome ^98^. Duplicate molecules were marked and removed using samtools ^99^. Reads with a quality >10 were binned into 500kb intervals and counted. IchorCNA was then used to perform GC correction and call the estimated tumor fraction using the following parameters: “—chrs “c(1,2,3,4,5,6,7,8,9,10,11,12,13,14,15,17,18,20,21,22) — normal “c(0.9,0.95,0.98)” –estimateScPrevalence FALSE –scStates “c()” –maxCN 3 – ploidy “c(2)” –normalPanel 20240722.CSF_PON_median.rds”. The panel of normal was based on 8 CSF sample without the presence of cancer and were not used in the study. Note: chr16 and chr19 were excluded due to the high variance on each of chromosomes. We and others have reported that increased GC content on these chromosome can produce higher numbers of false positives^100^. Next, we wanted to generate chromosome level calls. However, since ichorCNA only produces chromosome level not chromosome arm level calls, we then generated arm level calls using the median across each the GC corrected ichorCNA 500kb intervals. We discarded chr6p which produced numerous false calls associated with alignment artifacts surrounding the MHC regions.

### The “M” component of CSF-BAM (mutations)

Following library creation, two separate PCRs were designed to selectively enrich the Watson or Crick strand. Both PCRs used the same gene-specific primer, but each used a different anchoring primer. PCR duplicates derived from each strand could be distinguished by the orientation of the insert relative to the exogenous UID. Sequencing reads underwent initial processing by extracting the first 14 nucleotides as the exogenous barcode sequence (UIDs) and masking adapter sequencing Picard’s IlluminaBasecallsToSam (http://broadinstitute.github.io/picard). Reads were then mapped to the hg19 reference genome using BWA-MEM^98^ and sorted by barcode sequence using Samtools^99^. Duplex mutations were defined as mutations present in >80% of both the Watson and Crick families with the same UID. We had several metrics for the interpretation of mutations: only genomic positions with at least 2 or more reported annotations in the Catalogue of Somatic Mutations in Cancer (COSMIC) in genome-wide studies and were confirmed somatic mutations were considered^101^; only positions with at least two observations of mutant molecules were considered; only positions with at least 5x coverage were considered; only positions at least 30 bp away from the end of the molecule were considered.

During the analysis, we noticed two major outliers: FBXW7 codon 505 and KRAS codon 61. The frequency of mutations at these two codons was much higher than expected. Upon closer inspection, every molecule with a mutant FBXW7 codon 505 also had mutations at codons 289, 299, 300, and 314. We then performed blat analysis to compare the observed matches to any non-human genome^71^. The observed sequence perfectly matched the bovine genome. We hypothesized that bovine-derived hemostatic agents frequently used in neurosurgery^72^ may have contributed minor amounts of DNA that amplified and incorrectly aligned to FBXW7. We also found KRAS codon 61 mutations in two patients with trigeminal neuralgia. The significance of these mutations is unknown. To ensure high specificity of the assay and given the low abundance of this particular mutation in CNS cancers, we excluded all mutations at this position in our study.

### Statistical analysis

Cohort sample size was not selected for statistical power but rather was based on sample availability. The posterior probability that the assay is the most and least sensitive (or specific) will be calculated using Bayesian methods and we report credible intervals for the sensitivity and specificity of CSF-BAM. A uniform Beta(1,1) prior distribution will be used and we assume the number of correct assays follows a binomial distribution. The prior distribution will be used based on the observed data. Therefore, the resulting posterior distribution is Beta(1+# of success, 1+# failures). Statistics were calculated using GraphPad Prism 10. Statistical tests are specified in figure legends.

## Data Availability

All data produced in the present work are contained in the manuscript

## Data availability

Data generated in this study are available in the paper and supplementary files. Sequence data will be publicly available in the European Genome-phenome Archive prior to publication.

## Supplementary materials

Supplementary Table S1: Cohorts for CSF-BAM components

Supplementary Table S2A: SafeBSeqS primer sequences

Supplementary Table S2B: SafeTSeqS primer sequences

Supplementary Table S2B: Aneuploidy analysis primer sequences

Supplementary Table S2B: Mutation analysis primer sequences

Supplementary Table S3: Summary of SafeBSeqS analysis in WBC control samples

Supplementary Table S4: Summary of CSF-BAM SafeBSeqS analysis in CSF

Supplementary Table S5: Summary of SafeTSeqS analysis in WBC control samples

Supplementary Table S6: Summary of CSF-BAM SafeTSeqS analysis in CSF

Supplementary Table S7: Summary of aneuploidy analysis in control samples

Supplementary Table S8: Summary of CSF-BAM aneuploidy analysis in CSF samples

Supplementary Table S9: Reproducibility of aneuploidy analysis

Supplementary Table S10: Comparison of aneuploidy analysis with CSF-BAM and Real-CSF

Supplementary Table S11: CSF-BAM mutation training set in non-cancer CSF samples

Supplementary Table S12: CSF-BAM mutation analysis in CSF samples

Supplementary Table S13: Demographic characteristics by sample

Supplementary Table S14: Diagnostic categories by sample

Supplementary Table S15: CSF-BAM results summary

Supplementary Table S16: CSF-BAM detection based on CSF reservoir abutment by sample

Supplementary Table S17A: TCR clusters obtained via CSF-BAM

Supplementary Table S17B: TCR specificity annotation

Supplementary Figure S1: SafeBSeqS amplification

Supplementary Figure S2: SafeTSeqS amplification

Supplementary Figure S3: Aneuploidy reproducibility from two independent aliquots and libraries.

Supplementary Figure S4: Metrics for targeted mutation panel.

Supplementary Figure S5: Correlation of predicted aneuploidy neoplastic content to mutation neoplastic content.

Supplementary Figure S6: TCR UIDs and BCR UIDS recovered for each sample.

Supplementary Figure S7: T cell clonality and B cell clonality for evaluable samples with total UIDs ≥20.

Supplementary Figure S8: BCR clonality ROC and BCR IGHV4-34 gene segment usage

Supplementary Figure S9: Case reports demonstrating CSF-BAM clinical applicability

## Author contributions

Conceptualization: A.H.P., Y.W., B.V., C.D., C.B.

Data curation: A.H.P., Y.W., A.K., M.Parker., J.R.T., Y.X, R.G., M.A.H., J.T., C.D., C.B.

Formal analysis: A.H.P., Y.W., K.W.K., N.P., B.V., C.D., C.B.

Investigation: A.H.P., Y.W., J.D.C., J.D., L.D., M.Popoli., J.P., N.S., K.J., C.D.

Methodology: A.H.P., Y.W., J.D.C., K.W.K., N.P., B.V., C.D., C.B.

Resources: M.G., C.M.J., E.M.J., G.I.J., M. Lim, M. Luciano, D.M., J.N., S.R., C.H.S.,

J.W., C.K., A.M., M.G., D.K., K.C.S., C.A.P, M.H., S.P., C.B.

Writing – original draft: A.H.P., Y.W., B.V., C.D., C.B.

Writing – review & editing: A.H.P., Y.W., A.K., M.Parker., J.D.C., J.D., J.R.T., Y.X., R.G., M.A.H., J.T., L.D., M.Popoli., J.P., N.S., K.J., M.G., C.M.J., E.M.J., G.I.J., M. Lim, M. Luciano, D.M., J.N., S.R., C.H.S., J. W., C.K., A.M., M.G., D.K., K.C.S., C.A.P., M.H., S.P., K.W.K., N.P., B.V., C.D., C.B.

## Acknowledgements

We thank Cherie Blair for technical assistance and Elizabeth Cook for illustrations.

## Funding

Alex’s Lemonade Stand Foundation (C. B.)

American Society of Hematology Scholar award (S.P.)

Benjamin Baker Endowment 80049589 (C.D., Y.W.)

Burroughs Wellcome Career Award for Medical Scientists (C. B.)

Commonwealth Fund (C.B., N.P.)

Conrad R. Hilton Foundation (K.W.K., N.P., B.V.)

DOD award W81XWH-21-1-0251 (K.C.S.)

Doris Duke Career Development Award (K.C.S.)

Jerome Greene Foundation (C.H.S.)

Leukemia Lymphoma Society Translation Research Program award (S.P.)

Lustgarten Foundation for Pancreatic Cancer Research (B.V.)

NIH grant CA06973 (K.W.K., N.P., B.V.)

NIH grant DRP80057309 (C.D.)

NIH grant K08CA270403 (S.P.)

NIH grant RA37CA230400 (C. B.)

NIH grant R01CA276221 (C. B.)

NIH grant R21TR004059 (C. B.)

NIH grant T32GM136577 (J.D.C., A.H.P.)

NIH grant U01CA230691 (C. B., N.P.)

Sol Goldman Sequencing Facility at Johns Hopkins (B.V.)

Swim Across America Translational Cancer Research Award (C.D., S.P.)

Thomas M. Hohman Memorial Cancer Research Fund (C. B.)

Virginia and D.K. Ludwig Fund for Cancer Research (C. B., C.D., K.W.K., N.P., B.V.)

---

**Supplementary Figure S1.**
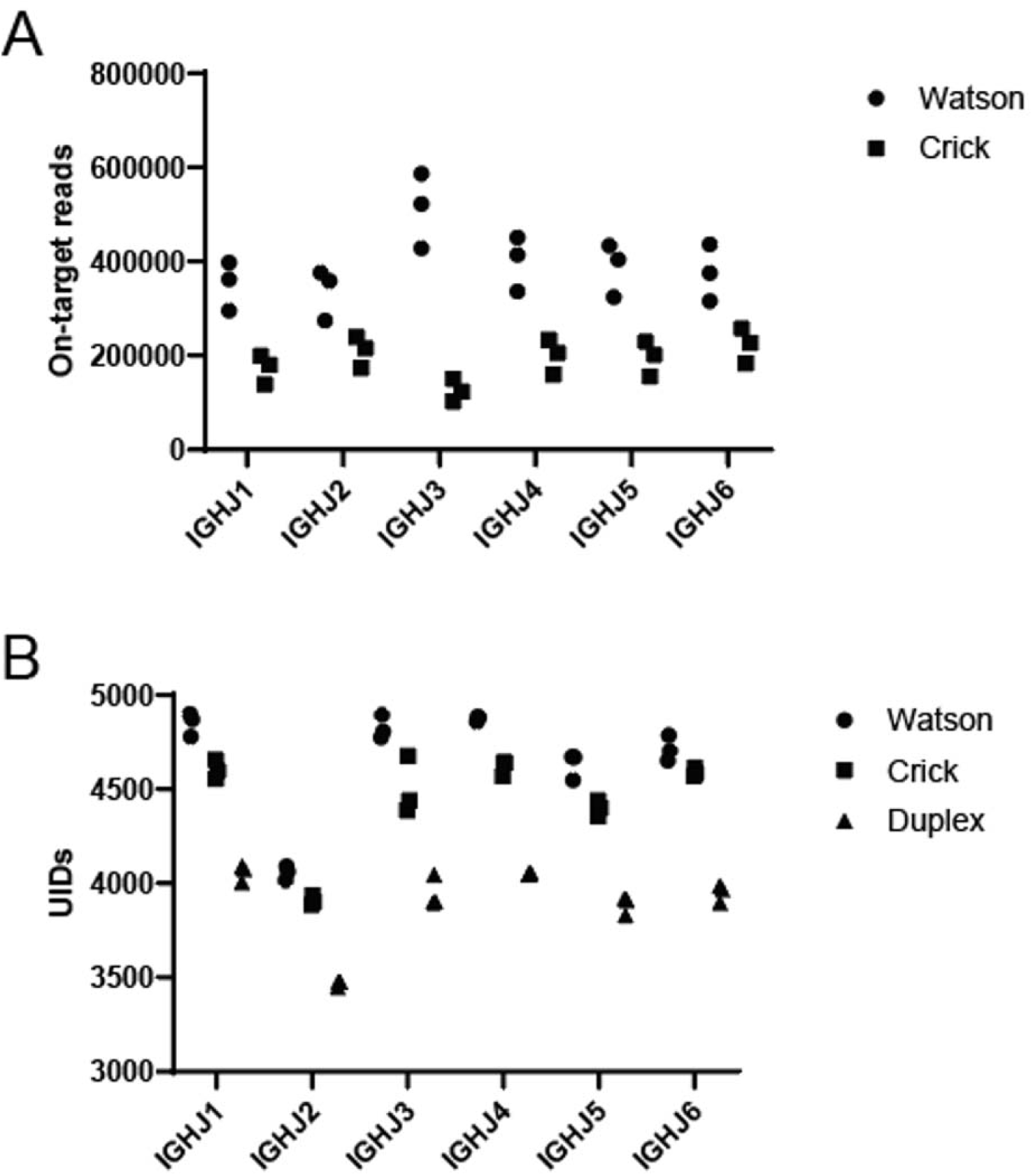
SafeBSeqS amplification. **A,** on-target reads and **B,** UIDs recovered for each IGHJ gene segment.

**Supplementary Figure S2.**
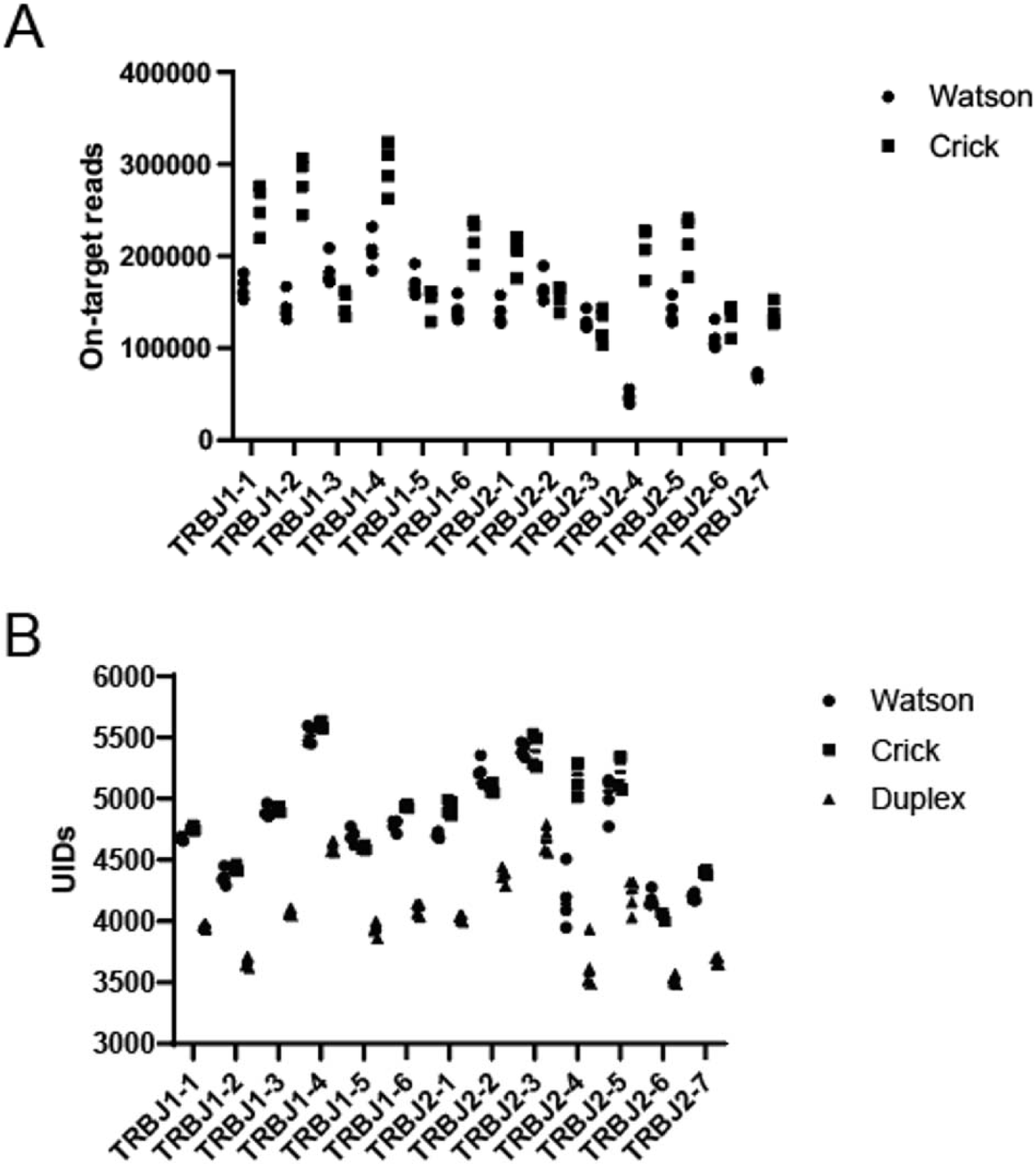
SafeTSeqS amplification. **A,** on-target reads and **B,** UIDs recovered for each TRBJ gene segment.

**Supplementary Figure S3.**
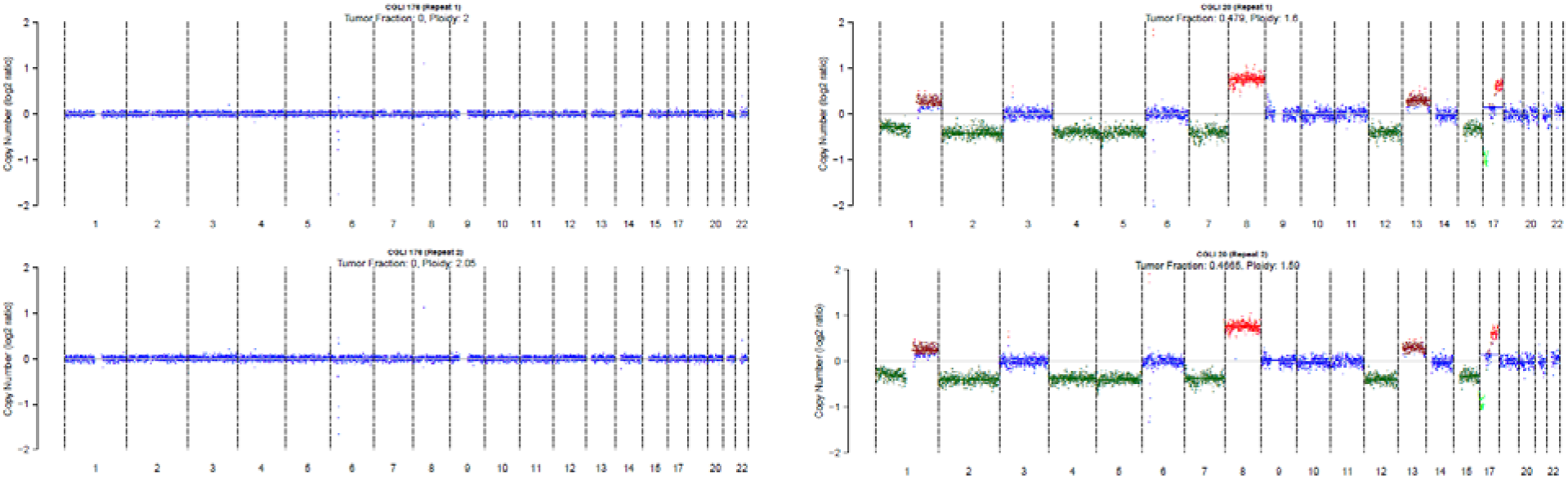
Aneuploidy reproducibility from two independent aliquots and libraries. **A,** A representative sample from a non-cancer individual that is predicted diploid in both replicates. **B,** A representative sample from an individual with cancer that is predicted aneuploid in both replicates.

**Supplementary Figure S4.**
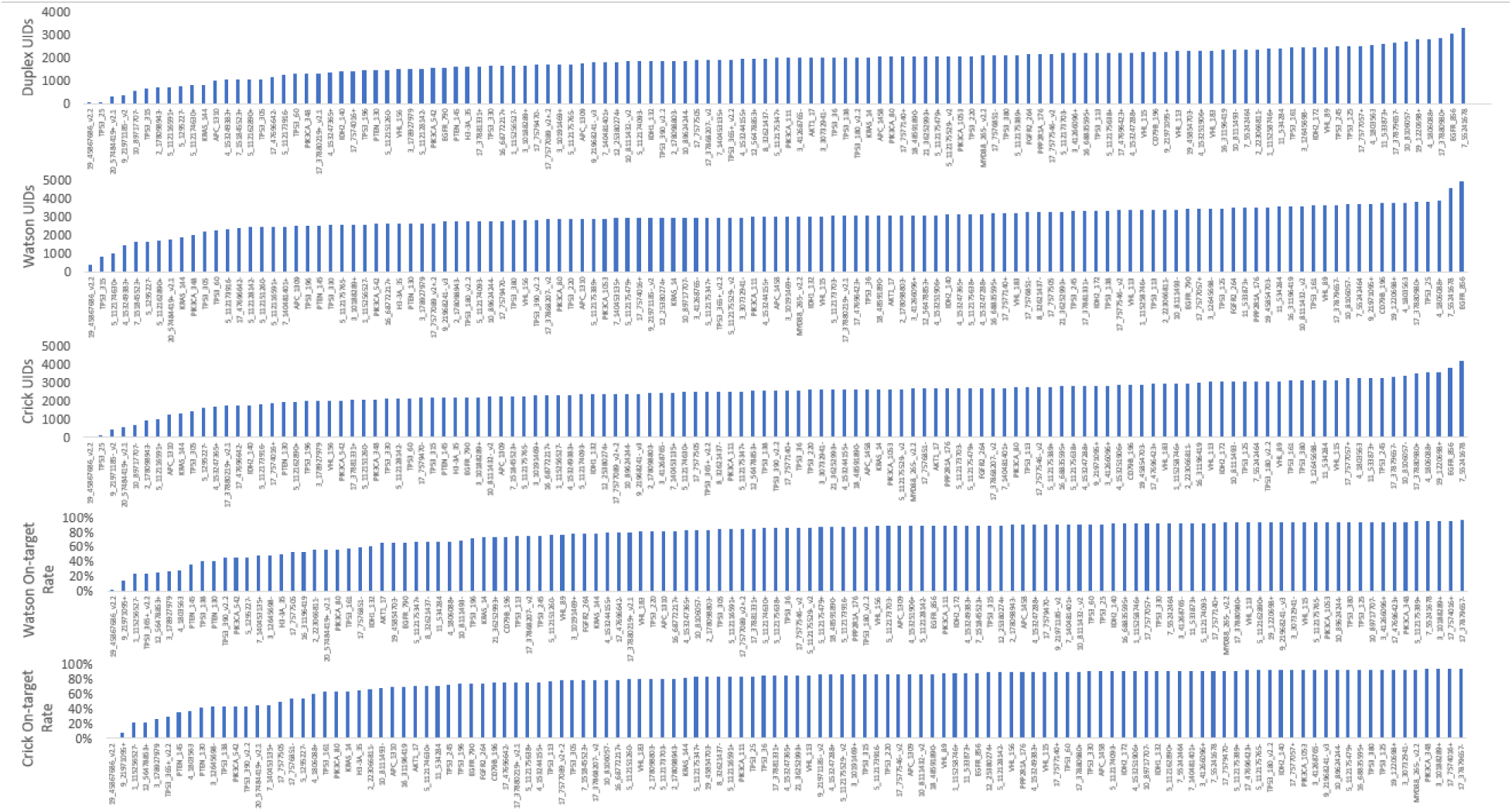
Metrics for targeted mutation panel. The number of duplex, Watson, and Crick UIDs, as well as Watson and Crick on-target rates are plotted.

**Supplementary Figure S5.**
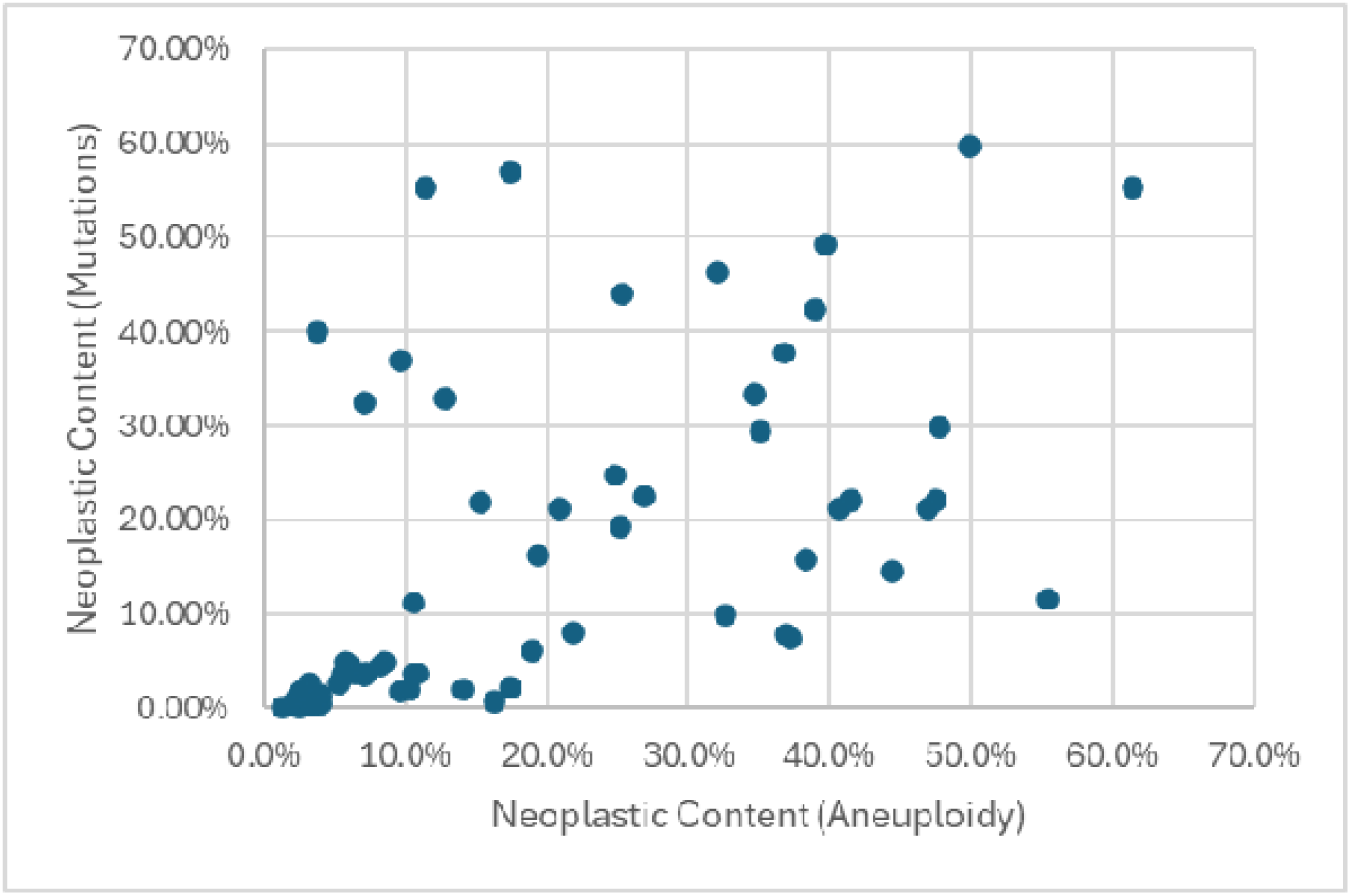
Correlation of predicted aneuploidy neoplastic content to mutation neoplastic content.

**Supplementary Figure S6.**
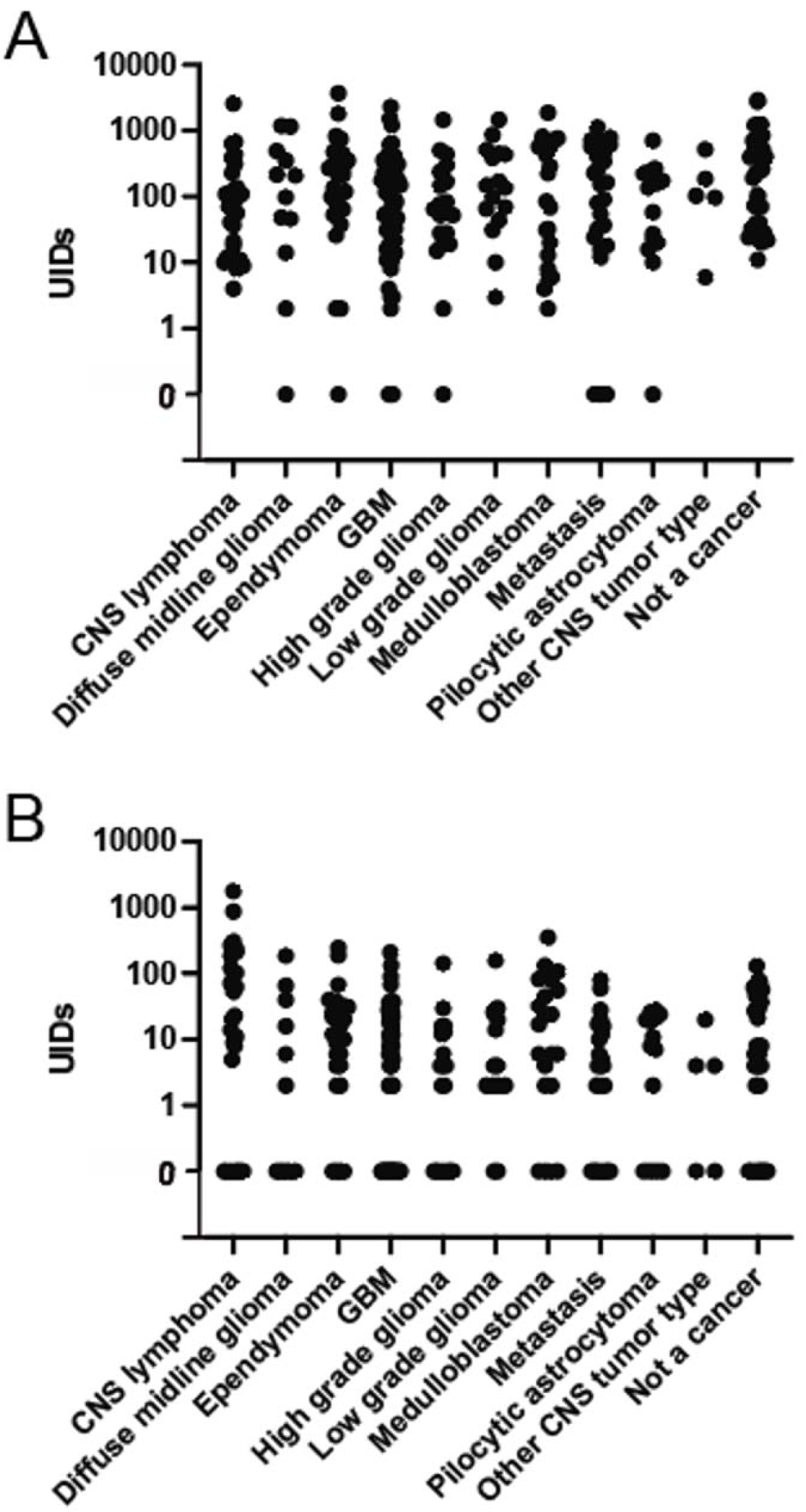
**A,** TCR UIDs and **B,** BCR UIDS recovered for each CSF sample.

**Supplementary Figure S7.**
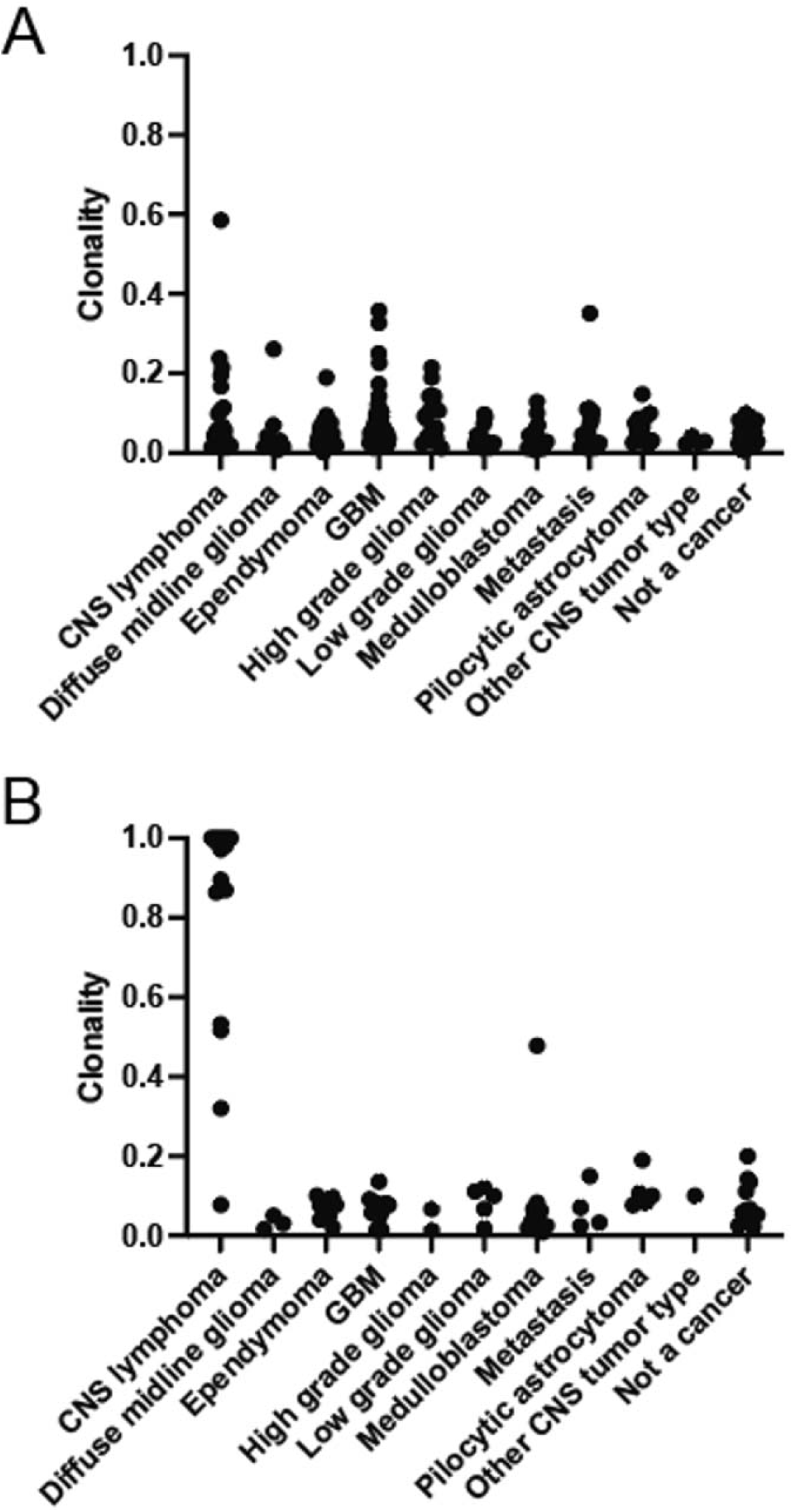
**A,** T cell clonality and **B,** B cell clonality for evaluable samples with total UIDs ≥20. B cell clonality variation P < 0.001 by Kruskal-Wallis test.

**Supplementary Figure S8.**
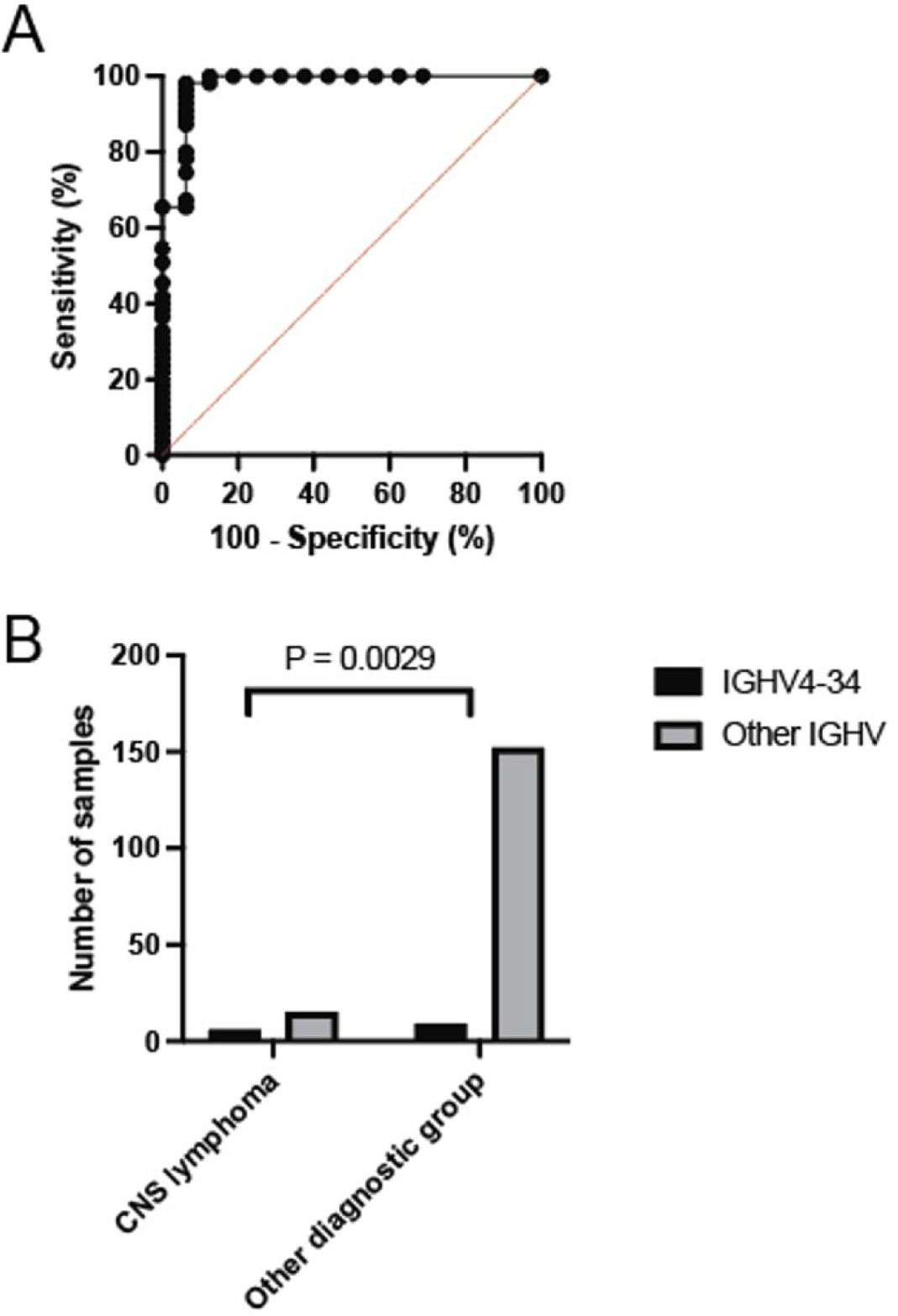
BCR clonality ROC and BCR IGHV4-34 gene segment usage. **A,** ROC for classification of CNS lymphoma samples versus samples of all other cancer types for evaluable samples with total UIDs ≥20. AUC = 0.98, 95% confidence interval 0.93-1.00. **B,** IGHV4-34 gene segment usage. Number of samples with the top clone using the IGHV4-34 gene segment versus all other IGHV segments for CNS lymphoma versus all other sample types. P = 0.0029 by Fisher’s exact test.

**Supplementary Figure S9.**
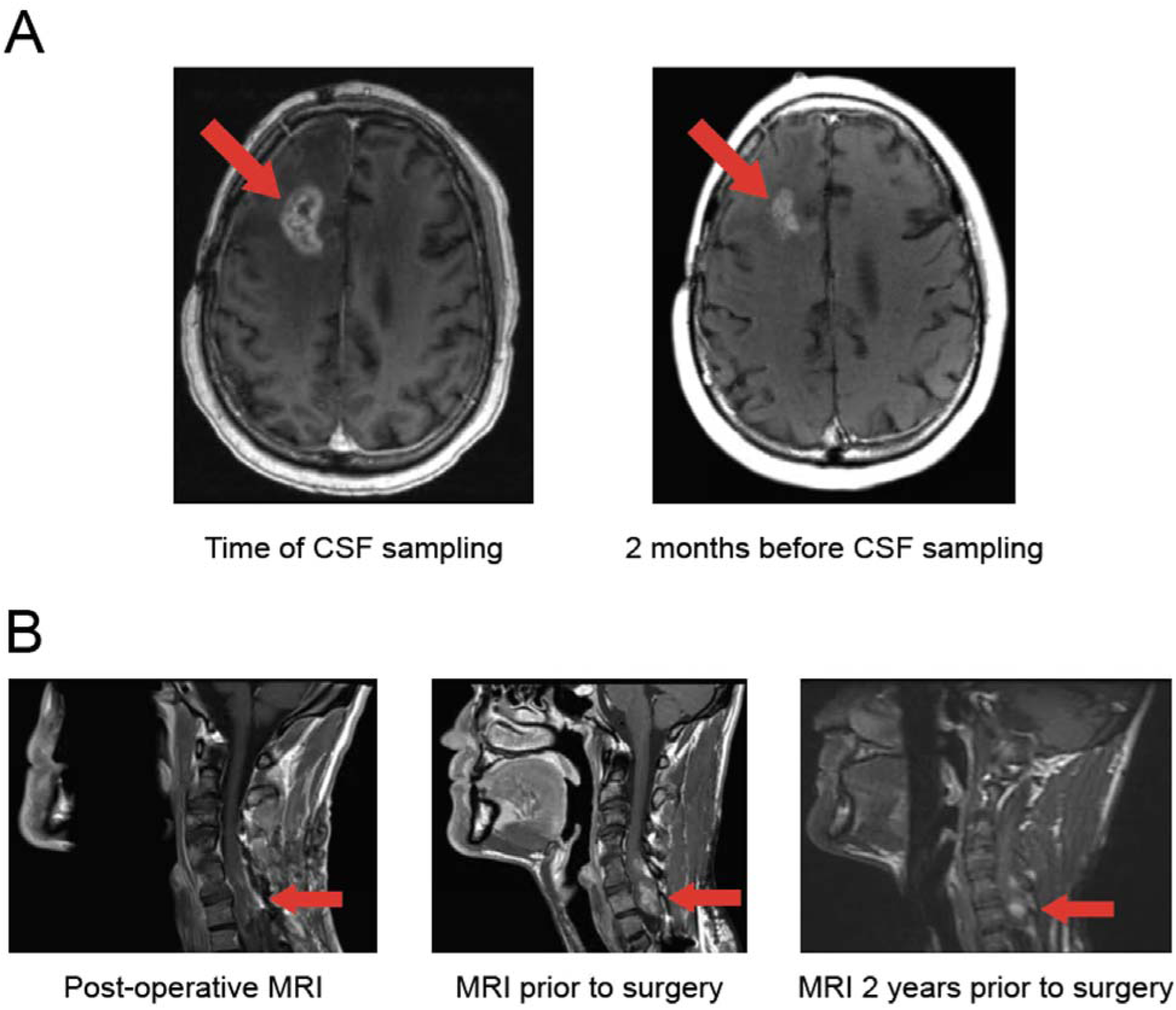
Case reports demonstrating CSF-BAM clinical applicability. **A,** CSF-BAM in the setting of pseudoprogression. Gadolinium enhanced axial MRI demonstrating increasing enhancement in right frontal lobe of patient GLIA793. The patient had previously been treated with chemotherapy and radiation therapy. Repeat resection demonstrated pseudoprogression and no active tumor. CSF-BAM was negative in a CSF sample obtained prior to the repeat resection where only pseudoprogression was detected. **B,** CSF-BAM can identify targetable mutations. Gadolinium enhanced sagittal MRI is shown demonstrating increasing size of a spinal ganglioglioma. CSF-BAM prior to resection detected a BRAF p.V600E mutation that can be targeted therapeutically.

